# Evaluating Conversational Image Segmentation for Medicine: Performance, Failure Modes, and a Fairness Audit Across Seven Modalities

**DOI:** 10.1101/2025.11.21.25340008

**Authors:** Justin Do, Nidhir Guggilla, Vivaswat Suresh, Rishi Kothari

## Abstract

**Introduction:** Medical-image segmentation underpins quantitative diagnostics and research, yet state-of-the-art models remain task-specific and data-hungry. The recent emergence of powerful, multimodal large language models (LLMs) presents a generalizable option; however, their efficacy in the specialized medical domain remains largely unquantified. We aim to benchmark the foundational Gemini 2.5 Flash and Gemini 2.5 Flash-Lite models for zero-shot medical image segmentation and evaluate them for potential bias in performance.

**Methods:** The models were tested on 13,086 medical images spanning seven distinct imaging modalities (endoscopy, fundoscopy, dermoscopy, laparoscopy, ultrasound, radiography, and CT) and 10 clinically relevant segmentation targets (e.g., colorectal polyps, skin lesions, liver tumors). Prompts followed a standard template (“Segment the … in this image.”). Per-image Dice and Intersection-over-Union (IoU) were computed against publicly released expert masks. Bias was assessed on 4,057 dermoscopy images split by Individual-Typology-Angle (ITA) into Light (> 28°, n = 3,499) and Dark (≤ 28°, n = 558) groups.

**Results:** Flash achieved a mean Dice=0.766 (IoU=0.680) for colorectal polyp segmentation, Dice=0.761 (IoU=0.672) for skin lesion segmentation, Dice=0.824 (IoU=0.736) for optic disc segmentation, and Dice=0.718 (IoU=0.616) for surgical tool segmentation, outperforming Flash-Lite by approximately 0.1 Dice points. Accuracy declined on low-contrast radiological tasks (Liver Mass CT Dice=0.071). In the fairness audit, Flash produced a successful mask for 2,839/3,499 light-tone images (81.2%) versus 378/558 dark-tone images (67.7%); χ² = 51.8, p < 0.001. Using all images, mean IoU=0.686 for light tones and IoU=0.591 for dark tones (Kruskal–Wallis H = 62.6, p < 0.001); Cliff’s δ = –0.208 (95% CI – 0.259 to –0.159).

**Discussion:** Gemini 2.5 Flash delivers competitive accuracy on high-contrast photographic datasets at negligible cost. Performance is weaker on radiographic modalities (ultrasound, CT, chest radiography), and in dermoscopy, we observe lower accuracy on darker ITA skin-tone groups. This study informs the field where foundational LLMs are deployment-ready for medical image segmentation and where targeted debiasing or domain adaptation is required.

## Introduction

Medical image segmentation enables pixel-level delineation of anatomy and pathology, supporting measurement, planning, and longitudinal response assessment. Beyond classification and bounding box detection, segmentation allows for characterization of diagnosis by providing an accurate measure of the size and shape of the target object, assisting in downstream tasks such as staging tumors. In cross-sectional imaging modalities such as CT/MRI, slice masks can be interpolated to produce 3D reconstructions for pre-procedural planning support.^1^ For the past decade, progress has been driven by task-specific machine learning models (U-Net, nnU-Net), which deliver strong accuracy but typically require curated labels, modality-specific preprocessing, and training or fine-tuning for each new target or domain. These demands constrain scalability and slow translation to clinical settings.^2,3^

Recently, foundational segmentation approaches have begun to achieve broader generalization. The Segment Anything Model (SAM) reframed segmentation as a promptable task, taking sparse spatial inputs (points, boxes, and masks) to produce object masks with zero-shot transfer on natural images. However, these approaches rely on spatial guidance, and their medical performance is uneven without adaptation.^4,5^ New advancements, such as language-conditioned methods, like CLIPSeg and LSeg, move closer to text-driven interaction by aligning image features with text embeddings, enabling masks from free-form labels, but they have seen limited clinical validation and are not designed for multi-turn, conversational refinement.^6,7^

Multimodal large language models have recently introduced a different interaction pattern: conversational segmentation. Gemini 2.5 exposes this capability through simple, natural-language prompts and returns structured outputs with per-mask bounding boxes and base-64 encoded PNG masks, eliminating the need for spatial prompting.^8^ To our knowledge, this work is among the first systematic evaluations in medical imaging of an LLM’s built-in conversational segmentation capability.

Beyond overall accuracy, another important consideration when evaluating a new machine learning method is equitable performance across demographic groups. Previous work has shown that machine learning vision models in chest X-ray classification tasks have exhibited underdiagnosis bias across multiple datasets, with systematically lower sensitivity in underserved groups.^9^ Compounding this risk, deep models can infer a patient’s race directly from medical images even after aggressive perturbations, implying that protected attributes (or their proxies) are embedded in visual data and may drive latent, differential errors.^10^ Dermatology is a particularly high-risk domain: darker skin tones are under-represented in common datasets, and state-of-the-art systems have shown reduced diagnostic performance on dark skin in external evaluations.^11,12^ Importantly for segmentation, recent work has documented measurable skin-tone bias in skin-lesion segmentation networks themselves, using standardized tone estimators such as the Individual Typology Angle (ITA).^13^ Large language models used in clinical contexts have also been shown to propagate race-based medical content and other demographic biases, emphasizing the need for detailed audits to characterize any performance differences between patient populations.^14^

In this work, we aim to (i) evaluate Gemini 2.5 Flash and Flash-Lite for zero-shot, natural-language segmentation across several medical modalities with task-level prompting, and (ii) test for skin-tone–linked disparities in dermoscopy using ITA groupings.

## Methods

### Image collection and tasks

To characterize Gemini’s segmentation performance across clinically relevant scenarios, we assembled 13,086 image–mask pairs spanning seven imaging modalities (endoscopy, fundoscopy, dermoscopy, laparoscopy, ultrasound, radiography, and CT) and 10 anatomical or pathological targets (polyp, optic disc, optic cup, surgical tool, uterus, breast lesion, pneumothorax, liver parenchyma, and liver tumor). We utilized publicly available machine learning datasets (Table 1): Kvasir-SEG, REFUGE 2, ISIC 2016, 2017, and 2018, AutoLaparo, BUSI, SIIM-ACR Pneumothorax, and LiTS 2017.^15–24^ This study falls under the common rule exception due to the use of publicly available, fully deidentified information.

**Table 1:** Images Evaluated for Segmentation.

| **Task (segmentation target)** | **Imaging modality** | **Images evaluated (n)** | **Source database** |
| --- | --- | --- | --- |
| Colonic polyp | Endoscopy (RGB colonoscopy) | 1000 | Hyper-Kvasir / Kvasir-SEG |
| Optic disc/cup | Color fundus photograph | 1200 | REFUGE2 |
| Skin lesion | Dermoscopy | 4074 | ISIC 2016, 2017, and 2018 Part 1*** |
| Surgical Tool and Uterus | Laparoscopy | 1786 | AutoLaparo |
| Breast lesion (benign + malignant) | Breast ultrasound | 647 | BUSI |
| Pneumothorax | Chest radiograph (AP/PA) | 2379* | SIIM-ACR Pneumothorax |
| Liver parenchyma | Abdominal CT | 1000** | LiTS 2017 |
| Liver tumor | Abdominal CT | 1000** | LiTS 2017 |

\*Only positive-case radiographs from the SIIM-ACR training set were included in the evaluation.
\*\*Random 1000-slice subsets of the full LiTS volumes were sampled.
\*\*\*ISIC databases were merged, and duplicate images were removed by comparing exact hash values.

### Segmentation Benchmarking

#### Image Pre-Processing

All source images exceeding 1024 × 1024 pixels were isotropically down-scaled to a maximum side length of 1024 pixels using bicubic resampling while preserving aspect ratio. Images at or below this dimension were retained at their native resolution and were not up-scaled. Gemini’s per-object segmentation masks, returned as base-64 encoded PNGs, were resized to their bounding-box extents with bilinear interpolation. Ground-truth masks supplied by each dataset were binarized using a high-luminance threshold appropriate to that dataset to create reference labels.

#### Model Inference

Segmentation was performed using Gemini 2.5 Flash and Flash-Lite with the documentation-recommended parameters. We set the *thinking budget* to 0 (disabling additional internal reasoning tokens to minimize run-to-run variance), used *temperature* = 0.5 (a mid-range setting that moderates randomness in token sampling), and set automated *safety filters* to Off to avoid false-positive blocking of benign anatomical terms. Each image received a single, task-level natural-language prompt (Supplement 1); model selection, parameters, and natural language prompt were all set based on Google’s published best practices for conversational image segmentation.^8^ Gemini’s per-pixel probability maps were converted to binary masks using the midpoint threshold (127/255 ≈ 0.5) after resizing each mask to its bounding box.^8,25^

#### Performance Metrics and Statistical Analysis

For every image, IoU (overlap divided by the union of predicted and reference areas; |intersection|/|union|) and Dice (2×overlap divided by the sum of areas; 2×|intersection|/[|prediction|+|reference|]) were computed between the binarized prediction and reference masks. Ninety-five-percent confidence intervals (95 % CIs) for mean IoU and mean Dice were estimated via non-parametric percentile bootstrapping with 1,000 replicates. Outputs were prospectively labeled as a false negative if no mask was returned.

#### Comparison Retrieval

We compare our results to an established segmentation baseline based on the performance of the Segment Anything Model on the same source datasets (Table 3).

**Table 2:** Performance Metrics for Various Segmentation Tasks.

| Task | Target | Model | Mean Dice (95% CI) | Mean IoU (95% CI) | FN Rate (%) |
| --- | --- | --- | --- | --- | --- |
| Colorectal Polyp | Polyp (N=1,000) | Gemini 2.5 Flash | 0.766 (0.748-0.782) | 0.680 (0.663-0.698) | 4.1% |
|  |  | Gemini 2.5 Flash-Lite | 0.684<br>(0.665-0.703) | 0.588<br>(0.570-0.606) | 6.8% |
| Skin Lesion<br>(ISIC) | Lesion<br>(N=4,074) | Gemini 2.5 Flash | 0.761<br>(0.753-0.769) | 0.672<br>(0.664-0.681) | 2.3% |
|  |  | Gemini 2.5 Flash-Lite | 0.716<br>(0.707-0.725) | 0.621<br>(0.612-0.629) | 2.1% |
| Retinal Fundus<br>(N=1,200) | Optic Disc | Gemini 2.5 Flash | 0.824<br>(0.812-0.835) | 0.736<br>(0.724-0.748) | 1.2% |
|  |  | Gemini 2.5 Flash-Lite | 0.360<br>(0.341-0.380) | 0.277<br>(0.262-0.294) | 0.9% |
|  | Optic Cup | Gemini 2.5 Flash | 0.240<br>(0.222-0.258) | 0.178<br>(0.166-0.193) | 56.3% |
|  |  | Gemini 2.5 Flash-Lite | 0.231<br>(0.217-0.245) | 0.156<br>(0.146-0.167) | 37.0% |
| Laparoscopy<br>(N=1,786) | Surgical Tool | Gemini 2.5 Flash | 0.718<br>(0.705-0.729) | 0.616<br>(0.603-0.628) | 1.1% |
|  |  | Gemini 2.5 Flash-Lite | 0.654<br>(0.642-0.666) | 0.536<br>(0.524-0.548) | 0.7% |
|  | Uterus | Gemini 2.5 Flash | 0.182<br>(0.168-0.195) | 0.142<br>(0.129-0.154) | 0.4% |
|  |  | Gemini 2.5 Flash-Lite | 0.303<br>(0.284-0.323) | 0.284<br>(0.263-0.305) | 30.3% |
| Breast US (BUSI) | Benign<br>(N=437) | Gemini 2.5 Flash | 0.501<br>(0.469-0.537) | 0.420<br>(0.388-0.454) | 1.1% |
|  |  | Gemini 2.5 Flash-Lite | 0.474<br>(0.445-0.501) | 0.366<br>(0.340-0.393) | 0.0% |
|  | Malignant<br>(N=210) | Gemini 2.5 Flash | 0.481<br>(0.439-0.521) | 0.368<br>(0.333-0.404) | 4.8% |
|  |  | Gemini 2.5 Flash-Lite | 0.487<br>(0.454-0.523) | 0.359<br>(0.331-0.390) | 0.0% |
| **Liver CT (LiTS)**<br>(N=1,000) | Liver | Gemini 2.5 Flash | 0.280<br>(0.260-0.299) | 0.212<br>(0.195-0.227) | 10.2% |
|  |  | Gemini 2.5 Flash-Lite | 0.189<br>(0.175-0.202) | 0.123<br>(0.114-0.133) | 3.6% |
|  | Liver Mass | Gemini 2.5 Flash | 0.071<br>(0.063-0.080) | 0.044<br>(0.038-0.050) | 5.2% |
|  |  | Gemini 2.5 Flash-Lite | --- | --- | --- |
| **Chest X-Ray**<br>(SIIM-ACR,<br>N=2,379) | Pneumothorax | Gemini 2.5 Flash | 0.035<br>(0.030-0.039) | 0.022<br>(0.019-0.025) | 0.7% |
|  |  | Gemini 2.5 Flash-Lite | 0.023<br>(0.020-0.026) | 0.013<br>(0.011-0.016) | 0.0% |

† Model did not return segmentations for this task due to safety policy alignment.

**Table 3:** Performance Compared to Baseline (SAM)

| Task | Dataset | Split / n | Setting notes (baseline) | Literature Dice (SAM) | Literature IoU (SAM) | Gemini 2.5 Flash Dice | Gemini 2.5 Flash IoU | Citations |
| --- | --- | --- | --- | --- | --- | --- | --- | --- |
| Colorectal polyp | Kvasir-SEG | full / 1,000 | SAM (base) 1-click point; | 0.663 | 0.496 | 0.766 | 0.680 | <sup>30</sup> |
| Skin lesion | ISIC 2017 | full / 2,594 | SAM: points | 0.6125 | — | 0.761 | 0.672 | <sup>31</sup> |
| Optic disc | REFUGE2 | full / 1,200 | SAM: point | 0.391 | — | 0.824 | 0.736 | <sup>32</sup> |
| Optic cup | REFUGE2 | full / 1,200 | As above | 0.335 | — | 0.240 | 0.178 | <sup>32</sup> |
| Breast mass \| benign | BUSI | full / 437 | SAM: Prompt unspecified | 0.5548 | 0.4550 | 0.501 | 0.420 | <sup>33</sup> |
| Breast mass \| malignant | BUSI | full / 210 | SAM: Prompt unspecified | 0.5548 | 0.4550 | 0.481 | 0.368 | <sup>33</sup> |
| Liver | LiTS | test / — | SAM: midpt prompted | 0.4354 | — | 0.280 | 0.212 | <sup>34</sup> |
| Liver tumor | LiTS | test / — | SAM: midpt prompted | 0.1442 | — | 0.071 | 0.044 | <sup>34</sup> |
| Pneumothorax | SIIM-ACR | test / — | No available literature | — | — | 0.035 | 0.022 | — |

**Table 4.**
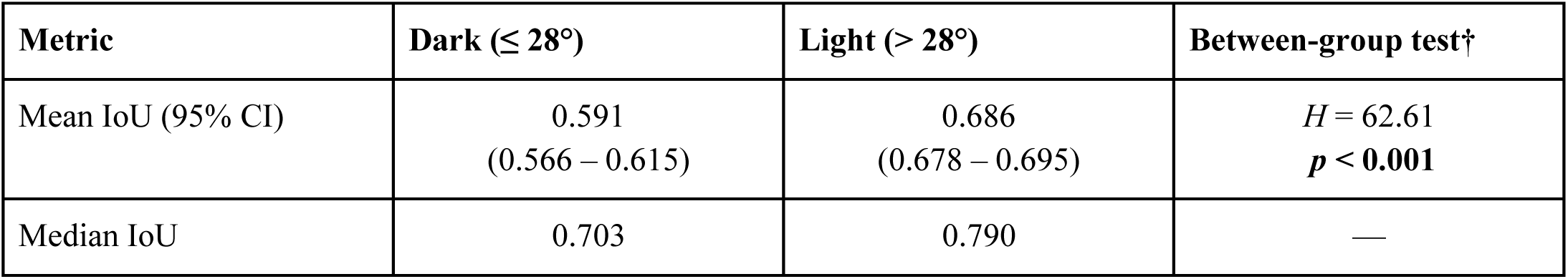
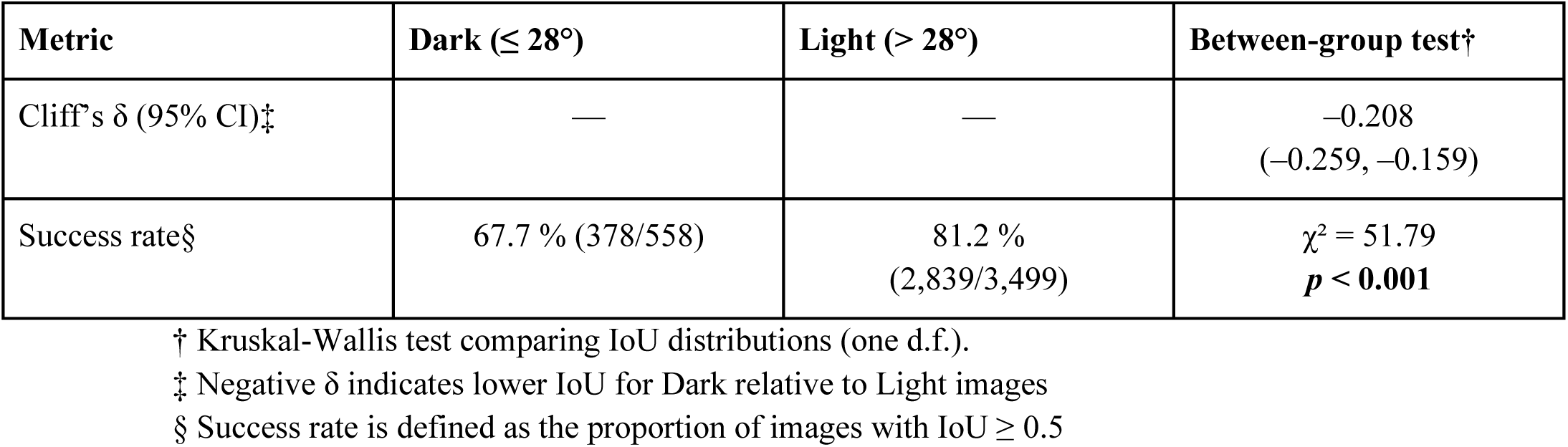
Gemini 2.5 Flash dermoscopy segmentation performance.

| Metric | Dark (≤ 28°) | Light (> 28°) | Between-group test† |
| --- | --- | --- | --- |
| Mean IoU (95% CI) | 0.591<br>(0.566 – 0.615) | 0.686<br>(0.678 – 0.695) | $H = 62.61$<br>$p < 0.001$ |
| Median IoU | 0.703 | 0.790 | — |

| Metric | Dark ($\leq 28^\circ$) | Light ($> 28^\circ$) | Between-group test† |
| --- | --- | --- | --- |
| Cliff's $\delta$ (95% CI)‡ | — | — | −0.208<br>(−0.259, −0.159) |
| Success rate§ | 67.7 % (378/558) | 81.2 %<br>(2,839/3,499) | $\chi^2 = 51.79$<br>$p < \mathbf{0.001}$ |

† Kruskal-Wallis test comparing IoU distributions (one d.f.).
‡ Negative $\delta$ indicates lower IoU for Dark relative to Light images
§ Success rate is defined as the proportion of images with IoU $\geq 0.5$

### Fairness Audit

We examine whether Gemini’s segmentation performance varied with skin tone. Each dermoscopy image underwent automated skin-tone labelling based on the Individual Typology Angle (ITA). ITA was calculated from the L* and b* channels of the CIE L*a*b* color space and expressed in degrees.^26^ All ITA classifications were confined to peri-lesional analysis obtained by inverting the ground-truth lesion mask.

Images were first converted to L*a*b*, after which candidate skin pixels were selected by a data-driven luminance window that retained values between a low-light floor and the 95th percentile, thereby preserving very dark skin while excluding specular highlights.^27^ We chose not to use a YCbCr skin filter due to recent literature that suggests it disproportionately filters Fitzpatrick V–VI tones in dermoscopic imagery.^28^ Images with less than two percent valid peri-lesional skin area or fewer than 200 candidate pixels were excluded to maintain ITA reliability.

For each retained pixel map, the pixel-wise ITA distribution was summarised by the median.^29^ Median ITA values were then mapped to the six-level Chardon typology and, for the primary fairness analysis, dichotomised into Light (> 28°) versus Dark (≤ 28°).^27^

For each skin-tone group, we reported the number of images (n) and the mean and median IoU. 95% Cis were obtained via bias-corrected and accelerated (BCa) bootstrapping with 5,000 resamples. Distributional differences were assessed using the Kruskal–Wallis H test (α = 0.05). When the global null was rejected, pairwise contrasts were performed with Dunn’s test and Holm–Bonferroni family-wise error control. Non-parametric effect sizes were quantified by Cliff’s Delta with percentile bootstrap confidence intervals. The proportion of “successful” segmentations (IoU ≥ 0.50) was compared across groups using a χ² test of independence.

## Results

Gemini 2.5 Flash performed variably across the benchmarks, with scores ranging from high in endoscopy (Polyp Dice=0.766), fundoscopy (OD Dice=0.824), dermoscopy (Lesion Dice=0.761), and laparoscopy (Surgical Tool Dice=0.718) to low in CT (Liver Mass Dice=0.071) and CXR (Pneumothorax Dice=0.035). Performance was strongest for blob-like, high-contrast structures, where Dice scores were high (up to 0.824) and fewer than 20% of cases fell below the 0.50 IoU low overlap threshold. In contrast, low-contrast thoracic and abdominal datasets (pneumothorax on chest radiograph, liver lesions on CT) proved challenging, with more than 60% of masks registering as outright misses or low IoU failures.

Across most datasets, Flash outperformed Flash-Lite, though there were notable exceptions (e.g., uterus and malignant breast lesion segmentation)

**Figure 1:**
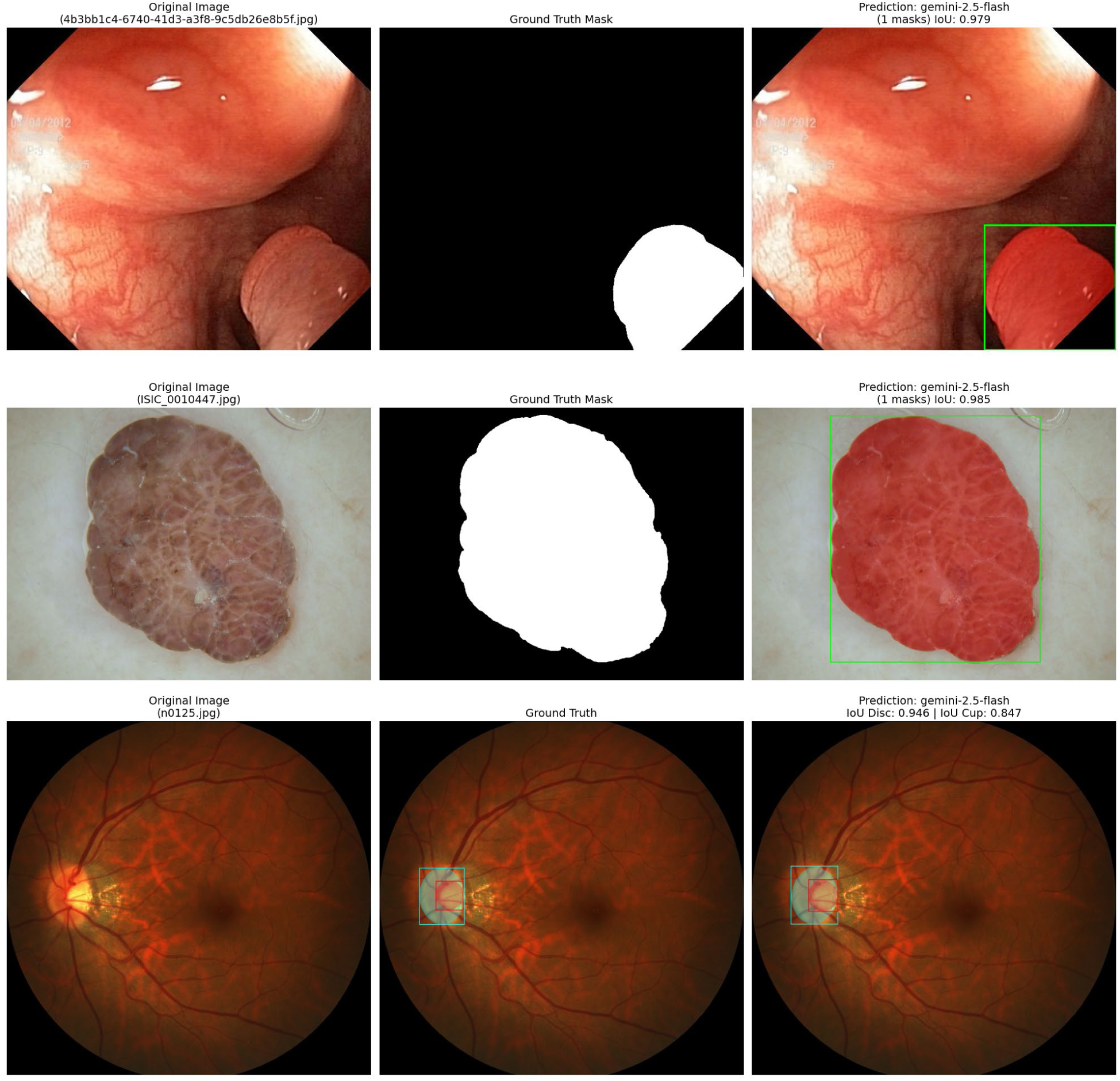
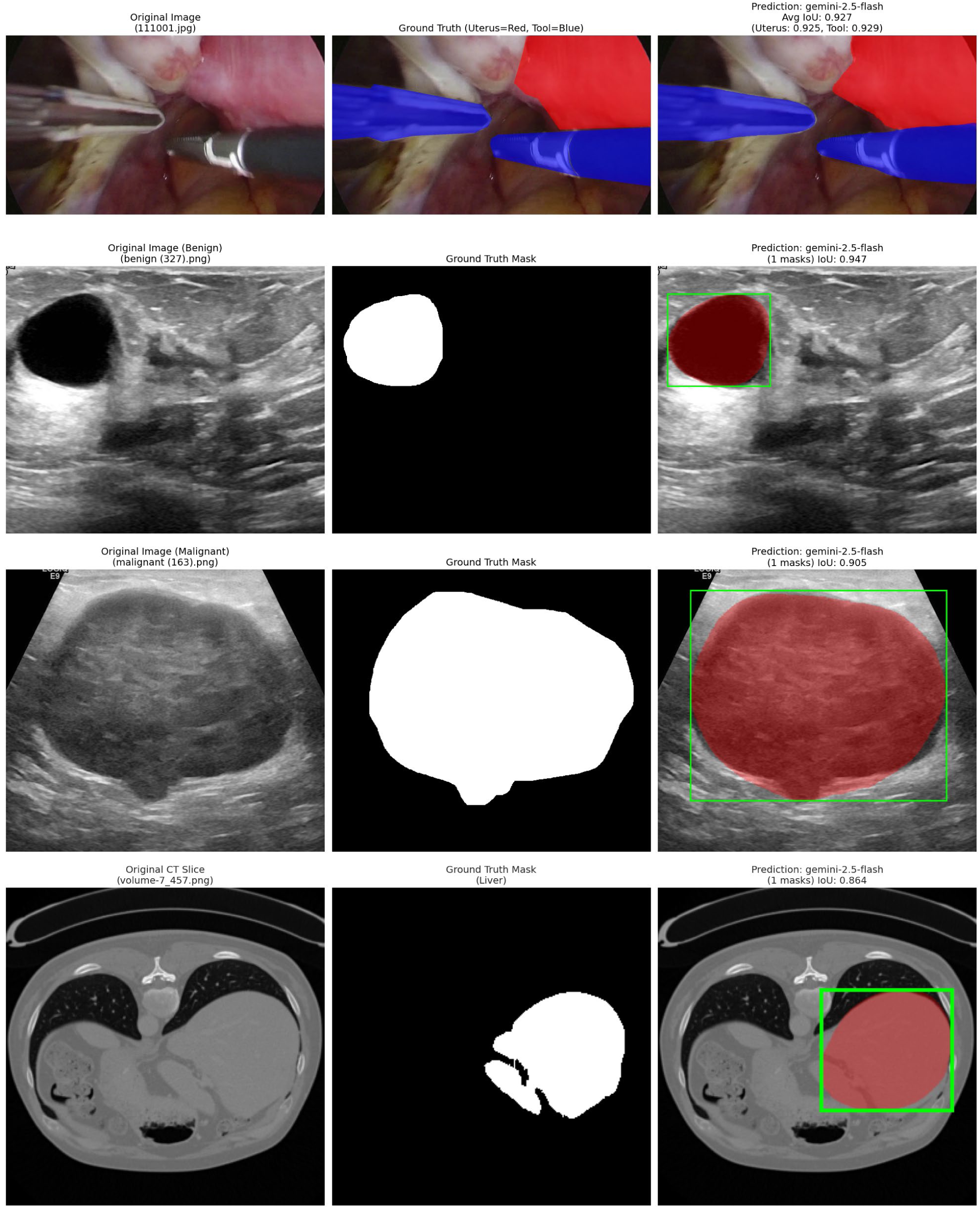
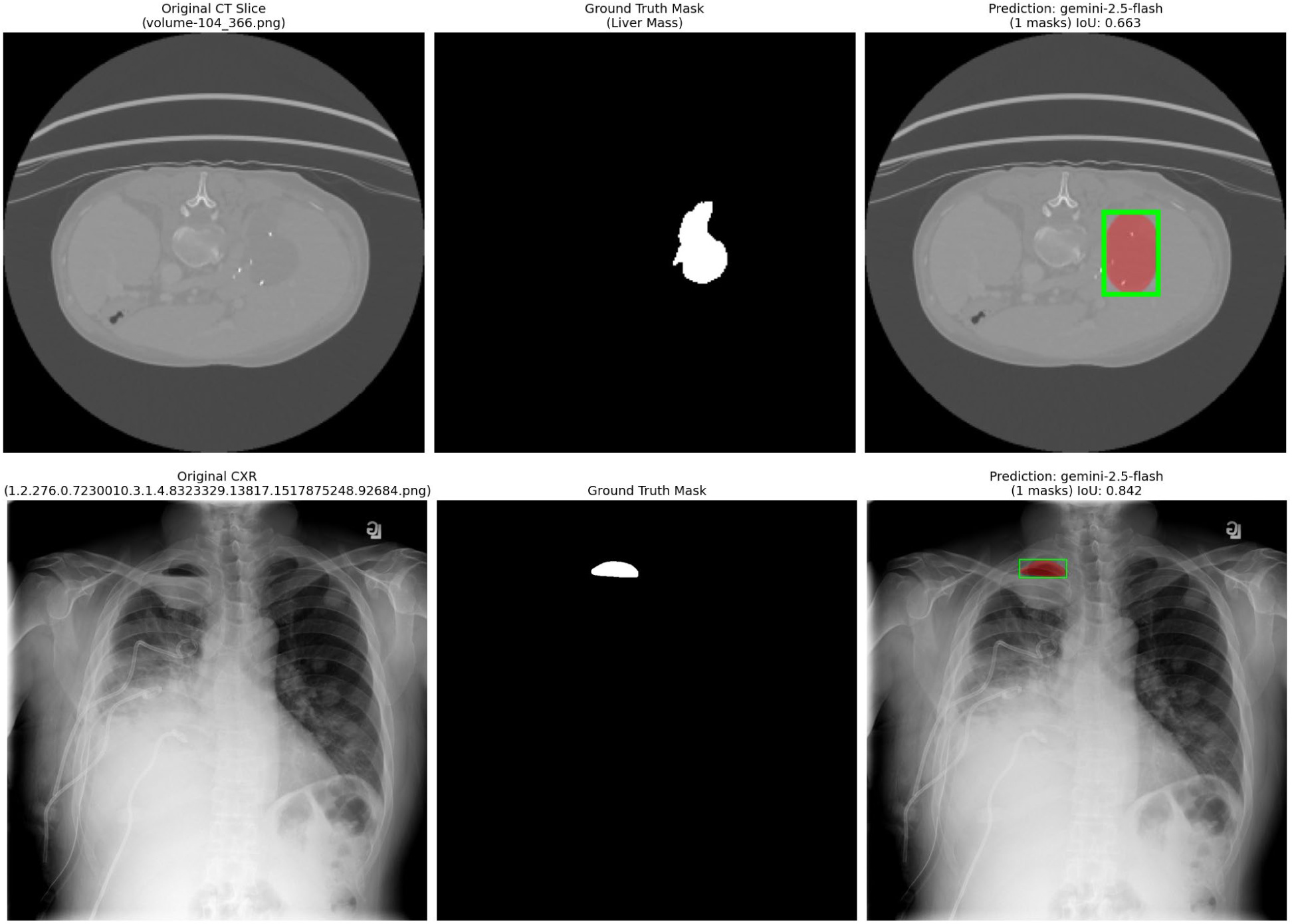
Best Case Segmentation by Gemini 2.5 Flash

### Fairness Audit

Among the 4,057 dermoscopy images evaluated by the fairness audit, 558 were classified as Dark (ITA ≤ 28°) and 3,499 as Light (ITA > 28°). In the full sample, Gemini 2.5 Flash achieved a mean IoU of 0.591 (95% CI 0.566–0.615) on dark-tone images versus 0.686 (0.678–0.695) on light-tone images (Kruskal-Wallis H = 62.61, p < 0.001); median IoU values were 0.703 and 0.790, respectively, with Cliff’s δ = - 0.208 (-0.259 to -0.159).

**Figure 2:**
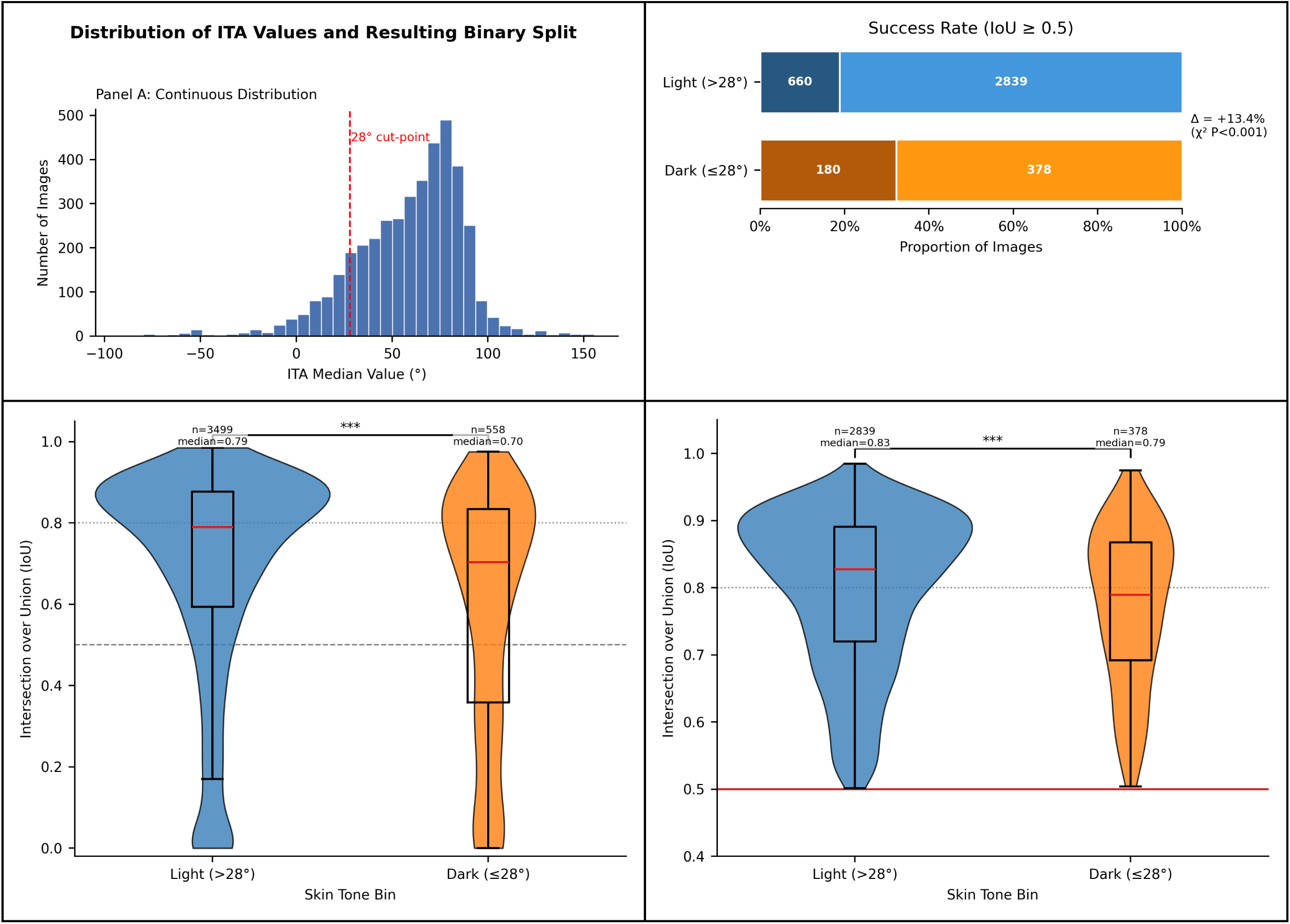
ITA Fairness Analysis

## Discussion

Our work demonstrates that large language models with conversational segmentation capabilities achieve zero-shot performance that is competitive with, and sometimes superior to, current spatially prompted generalist models, representing a new opportunity for medical image analysis. This performance varies significantly depending on the imaging modality and target characteristics. Furthermore, our fairness audit revealed performance disparities based on patient skin tone, underscoring the need for rigorous validation and targeted de-biasing to achieve equitable clinical machine learning.

### High Performance from Conversational Segmentation

Gemini’s ability to perform segmentation via natural language prompts improves upon prior models that required spatial input. For high-contrast targets like colorectal polyps, skin lesions, and optic discs, Gemini 2.5 Flash outperformed point-prompted Segment Anything Model (SAM) studies in mean Dice score (polyp: 0.766 vs. 0.663; lesion: 0.761 vs. 0.613; disc: 0.824 vs. 0.391) without manual guidance.^30–34^ This conversational interface significantly lowers the technical barrier for clinicians, a benefit supported by literature showing that other language-based tools, such as speech recognition, consistently reduce report turnaround times in radiology.^35^ Combined with a low API cost of approximately $0.001 per image, this approach could democratize access to advanced segmentation by removing dependencies on expensive hardware or local model deployment.

### Stratification of Performance by Image Contrast and Domain Familiarity

The model’s performance across our benchmark was clearly stratified by the visual characteristics of the segmentation task.

Gemini 2.5 Flash performed best on tasks involving well-defined, high-contrast targets in imaging modalities that resemble natural photographs, such as endoscopy, dermoscopy, fundoscopy, and laparoscopy. For colorectal polyps, skin lesions, optic discs, and laparoscopy, the model achieved high Dice scores (0.766, 0.761, 0.824, and 0.718, respectively) and low failure rates. These tasks share key features: a clear boundary between the target and surrounding tissue and a visual presentation similar to the general-domain images on which foundation models are primarily trained. This suggests the model can effectively be generalized from its presumably general ImageNet-style training images and applied to these “naturalistic” medical imaging tasks.

Performance degraded for targets with ambiguous or low-contrast boundaries. For optic cup segmentation, a task characterized by a subtle topographic shift rather than a clear edge, the model often failed to identify the target entirely, resulting in a false-negative rate of 56.3%. Similarly, for breast lesions on ultrasound, which frequently have ill-defined margins, the IoU scores revealed a bimodal distribution (Supplemental 2): a large peak of failures (IoU near 0) and a second peak of successful segmentations, with few cases in between.

Finally, the model performed worst on low-contrast tasks within specialized radiological modalities that are visually dissimilar from its general training data. For both liver parenchyma and liver tumor segmentation on abdominal CT, failure rates exceeded 60%. Performance was poorest for pneumothorax segmentation on chest radiographs, with a mean Dice score of just 0.035, indicating a near-complete failure to identify the target. This suggests the model’s foundational capabilities do not yet transfer to interpreting the abstract patterns of grayscale radiological imaging, likely due to a lack of domain-specific features learned during training.

### Model Capability and Failure Modes

Across nearly all tasks, Gemini 2.5 Flash consistently outperformed its smaller Flash-Lite counterpart. For many tasks, this resulted in a moderate decrease in segmentation quality, but the gap was most pronounced for optic disc segmentation. In this specific task, the drop in performance was catastrophic, with the mean Dice score falling from a clinically useful 0.824 with Flash to a non-functional 0.360 with Flash-Lite, demonstrating that model selection has a significant impact on clinical utility (Supplemental 3).

This performance hierarchy is compounded by features inherent to the model output. The bimodal IoU distributions observed in several tasks indicate that the models do not exhibit graceful degradation; instead, they tend to either produce an accurate mask or miss the target entirely, resulting in poor localization (Supplemental 4). Unlike traditional segmentation models that can output full-frame pixel-level probability maps, Gemini returns a probability mask only within its own predicted bounding box, decreasing its explainability outside of its predicted target localization.^5^ This combination of silent failures and opaque confidence levels may present a safety consideration, potentially leading to missed findings and increasing the cognitive load on the supervising clinician who must adjudicate these binary outcomes.^36^

### Bias in Segmentation Performance

Our fairness audit revealed a significant, systematic performance degradation when segmenting lesions on darker skin tones. Gemini 2.5 Flash produced a successful mask (IoU ≥ 0.50) for 81.2% of light-tone images but only 67.7% of dark-tone images, and the mean IoU was significantly lower for the dark-tone group (Cliff’s δ = -0.208). This performance gap is likely multifactorial, rooted in both unrepresentative training data and intrinsic technical challenges.

The disparity is likely driven, in part, by the composition of the public datasets used to train foundation models. Multiple audits of public dermatoscopy databases have confirmed a marked underrepresentation of images from individuals with darker skin tones (Fitzpatrick types IV-VI), with some estimates suggesting that over 90% of images are from fair-skinned individuals.^37,38^

Upstream inequities in clinical pathways likely contribute to downstream data imbalance. Large-scale experiments show that both dermatologists and primary-care physicians are less accurate in evaluating images of darker skin, with skin-tone–linked differences in triage decisions (e.g., biopsy recommendations) also observed.^39^ Concurrently, Black and Hispanic patients have lower outpatient access to dermatology (and longer wait times/acceptance barriers under Medicaid).^40^ In addition, dermoscopy imaging is predominantly captured in dermatology departments and remains uncommon in primary care. Together, these factors make it likely that fewer dermoscopic images are generated for darker-skinned patients to be used in dataset curation and model training.

This data imbalance is compounded by an intrinsic technical challenge. Low contrast between a lesion and the surrounding skin has long been recognized as a primary obstacle to the accurate automated detection of borders in dermoscopy.^41^ Recent fairness analyses confirm that segmentation performance correlates with skin color and that low lesion-skin contrast disproportionately degrades model performance on images from patients with higher-pigmentation skin.^13^ Our findings are therefore consistent with a dual-problem hypothesis: the model is undertrained on images of darker skin, and those images are often more technically challenging to segment.

Our findings are not unique to Gemini 2.5 Flash/Flash-Lite or image segmentation. Implicit and structural bias is often unexpectedly encoded in the data collected, from which foundational models are then trained. Across modalities, racial and SES bias has been identified in health risk scores, chest radiography classifiers, and LLMs.^10,42–44^ Our findings, along with the growing body of literature, emphasize the importance of practitioners and users of clinical machine learning critically examining data sources for bias and incorporating bias-mitigation strategies during data collection and model training.

### Limitations

This study has several limitations. First, our evaluation relied on publicly available, research-grade datasets, which may not reflect the heterogeneity of imaging devices, acquisition protocols, and patient populations encountered in real-world clinical practice. Second, our baseline comparisons to SAM were drawn from published literature rather than direct, head-to-head runs on identical data splits, which introduces a potential source of variability. Third, our methodology employed a static, standardized prompt for each task and did not explore the potential for iterative, conversational prompting to refine or rescue initial failures. Fourth, for volumetric modalities like CT, our 2D, per-slice analysis lacks the full 3D context that a radiologist would typically use, likely contributing to the poor performance on these tasks. Finally, while safety filters were disabled, some model-task combinations resulted in API refusals, an opaque failure mode that limits real-world reliability.

### Future Work

The limitations of this study highlight several key avenues for future research. To address heterogeneous results, future studies should consider running Med-SAM or SAM against the same corpus of image-mask pairs with zero-shot prompting adapters. To address performance gaps, future work should explore advanced prompting techniques, such as multi-turn conversational refinement or few-shot examples, and test hybrid workflows where the language model provides an initial region of interest to a specialized segmentation model. Most critically, the fairness deficits identified here must be addressed through the curation of more demographically representative training datasets and the employment of technical solutions, such as weighted loss functions, to ensure equitable performance.

In conclusion, conversational segmentation using general-purpose foundation models is a novel and highly accessible paradigm for medical image analysis. Our results indicate that this approach is viable today for specific high-contrast tasks and demonstrates considerable potential for broader application. The primary obstacles to widespread adoption are the performance variability across different imaging modalities and the significant algorithmic biases we identified. Future iterations of these models, particularly those incorporating domain-specific fine-tuning and more representative training data, are expected to mitigate these limitations, leading to more robust and equitable performance.

## Supporting information

Multimedia Supplemental

## Data Availability

All data produced in the present study are available upon reasonable request to the authors

https://github.com/DebeshJha/Kvasir-SEG

https://www.kaggle.com/datasets/victorlemosml/refuge2

https://challenge.isic-archive.com/data/

https://autolaparo.github.io/

https://www.kaggle.com/datasets/sabahesaraki/breast-ultrasound-images-dataset

https://www.kaggle.com/datasets/jesperdramsch/siim-acr-pneumothorax-segmentation-data

https://www.kaggle.com/datasets/andrewmvd/lits-png

