## Supplementary material for "Evaluating Conversational Image Segmentation for Medicine: Performance, Failure Modes, and a Fairness Audit Across Seven Modalities": Multimedia Supplemental

### Supplemental 1 - Prompt and API Settings

To ensure consistent segmentation performance on all tasks, we adapted the best practices for prompting Gemini 2.5 models for segmentation:

#### API Settings

temperature = 0.5
thinking_budget = 0 (disabled)
types.SafetySetting(category=types.HarmCategory.HARM_CATEGORY_HARASSMENT, threshold=types.HarmBlockThreshold.BLOCK_NONE),
types.SafetySetting(category=types.HarmCategory.HARM_CATEGORY_HATE_SPEECH, threshold=types.HarmBlockThreshold.BLOCK_NONE),
types.SafetySetting(category=types.HarmCategory.HARM_CATEGORY_SEXUALLY_EXPLICIT, threshold=types.HarmBlockThreshold.BLOCK_NONE),
types.SafetySetting(category=types.HarmCategory.HARM_CATEGORY_DANGEROUS_CONTENT, threshold=types.HarmBlockThreshold.BLOCK_NONE),

#### Prompts

##### Polyps

Give the segmentation masks for colorectal polyp. Output a JSON list of segmentation masks where each entry contains the 2D bounding box in the key "box_2d", the segmentation mask in key "mask", and the text label in the key "label". Use descriptive labels.

##### Skin Lesion

Give the segmentation masks for all skin lesions. Output a JSON list of segmentation masks where each entry contains the 2D bounding box in the key "box_2d", the segmentation mask in key "mask", and the text label in the key "label".

##### Optic Disc/Cup

Give the segmentation masks for the optic disc and the optic cup in this retinal fundus image. Output a JSON list of two segmentation masks. One entry should have the label "optic disc" and the other "optic cup". Each entry must contain the 2D bounding box in "box_2d", the segmentation mask in "mask", and the text label in "label".

#### Surgical Tool/Uterus

In this video frame from a laparoscopic hysterectomy, provide segmentation masks for the anatomy and surgical tools. Output a JSON list of segmentation masks. - Use the label "uterus" for the uterus. - Use the label "surgical tool" for any and all surgical instruments, including grasping forceps, LigaSure, hooks, and their shafts and manipulators. Combine all instrument parts into one category. Each JSON entry must contain the 2D bounding box in the key "box_2d", the segmentation mask in key "mask", and the text label in the key "label".

#### Breast Mass

Give the segmentation mask for the breast mass in this ultrasound image. Output a JSON list of segmentation masks where each entry contains the 2D bounding box in the key "box_2d", the segmentation mask in key "mask", and the text label in the key "label". Use a descriptive label like "mass".

#### Liver/Liver Mass

Where {SEGMENTATION_TARGET} is “liver” or “liver mass”
In this slice of an abdominal CT, give the segmentation mask for the {SEGMENTATION_TARGET}. Output a JSON list of segmentation masks where each entry contains the 2D bounding box in the key "box_2d", the segmentation mask in key "mask", and the text label in the key "label". Use the label "{SEGMENTATION_TARGET}".

#### Pneumothorax

In this chest x-ray, give the segmentation mask for the pneumothorax. Output a JSON list of segmentation masks where each entry contains the 2D bounding box in the key "box_2d", the segmentation mask in key "mask", and the text label in the key "label". Use the label "pneumothorax".

### Supplemental 2 - BUSI Segmention IoU Plot

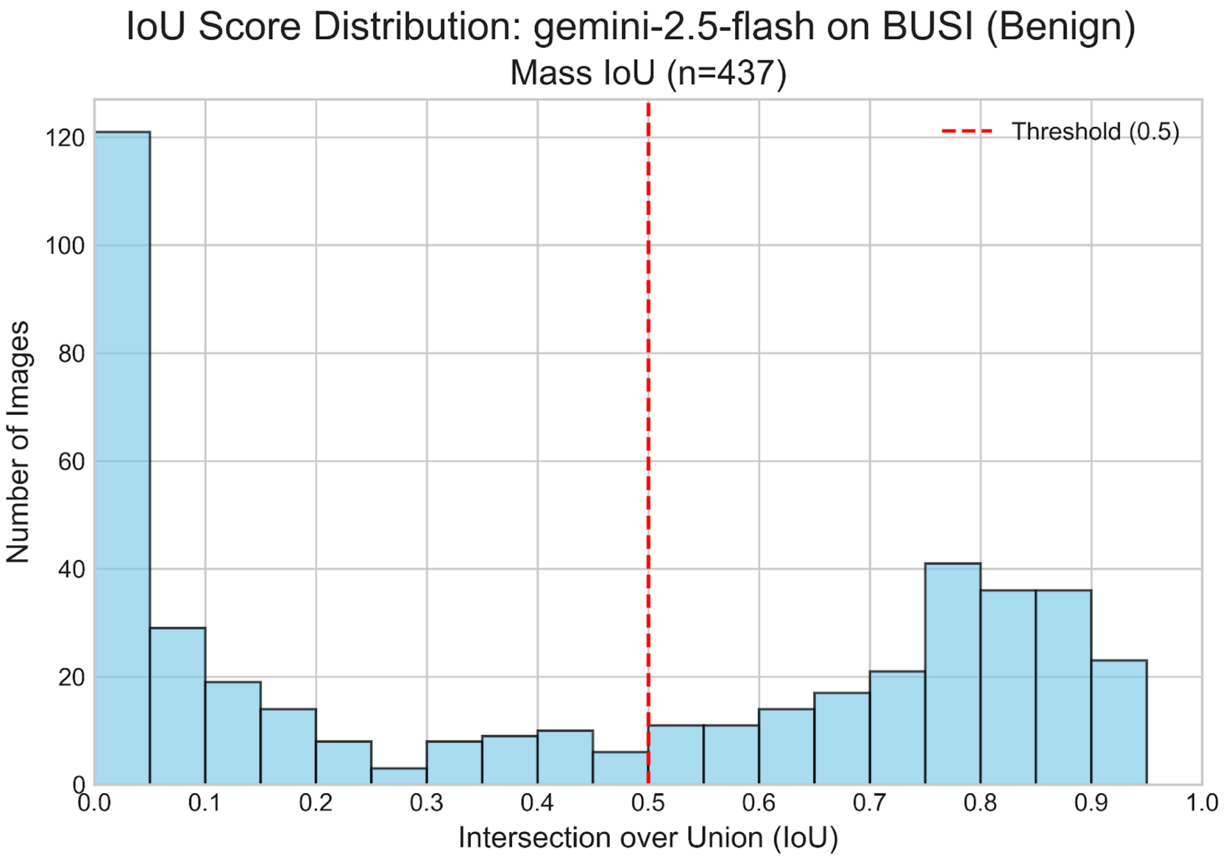

### Supplemental 3 - Optic Disc IoU Plots Flash vs. Flash-Lite

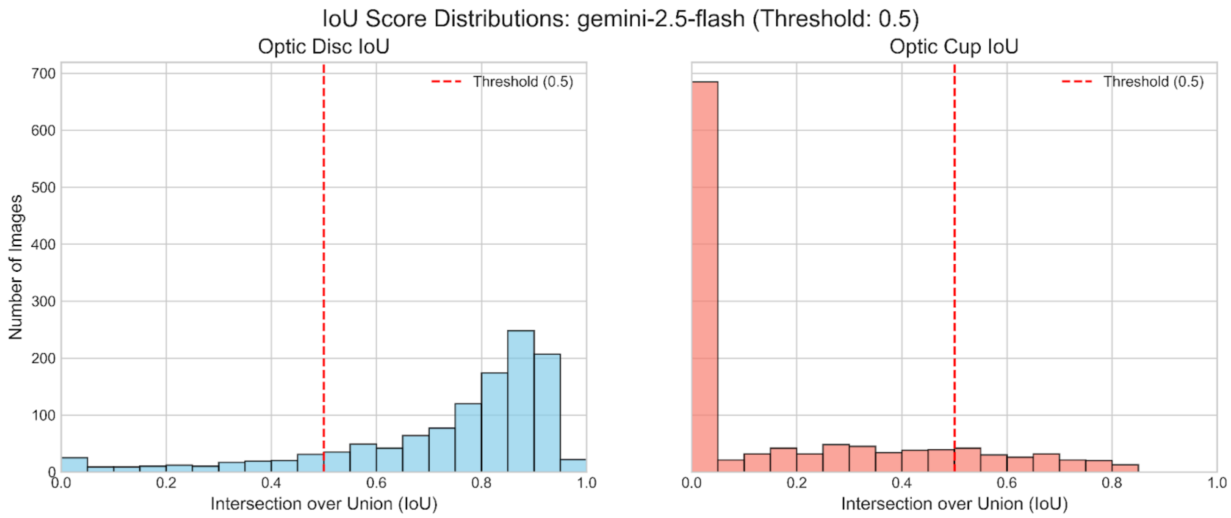


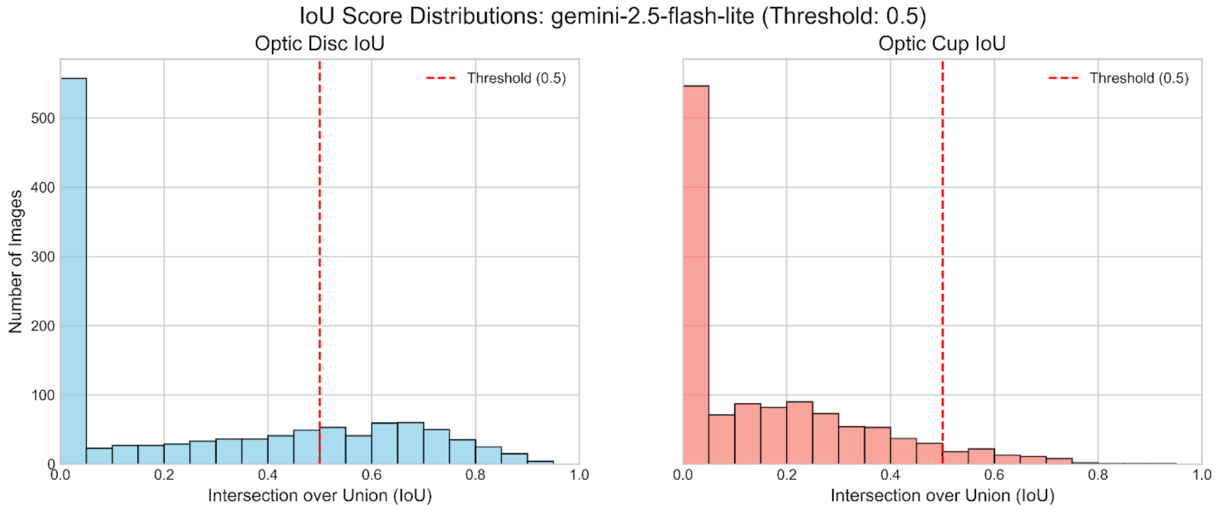

### Supplemental 4 - Skin Lesion IoU Plot

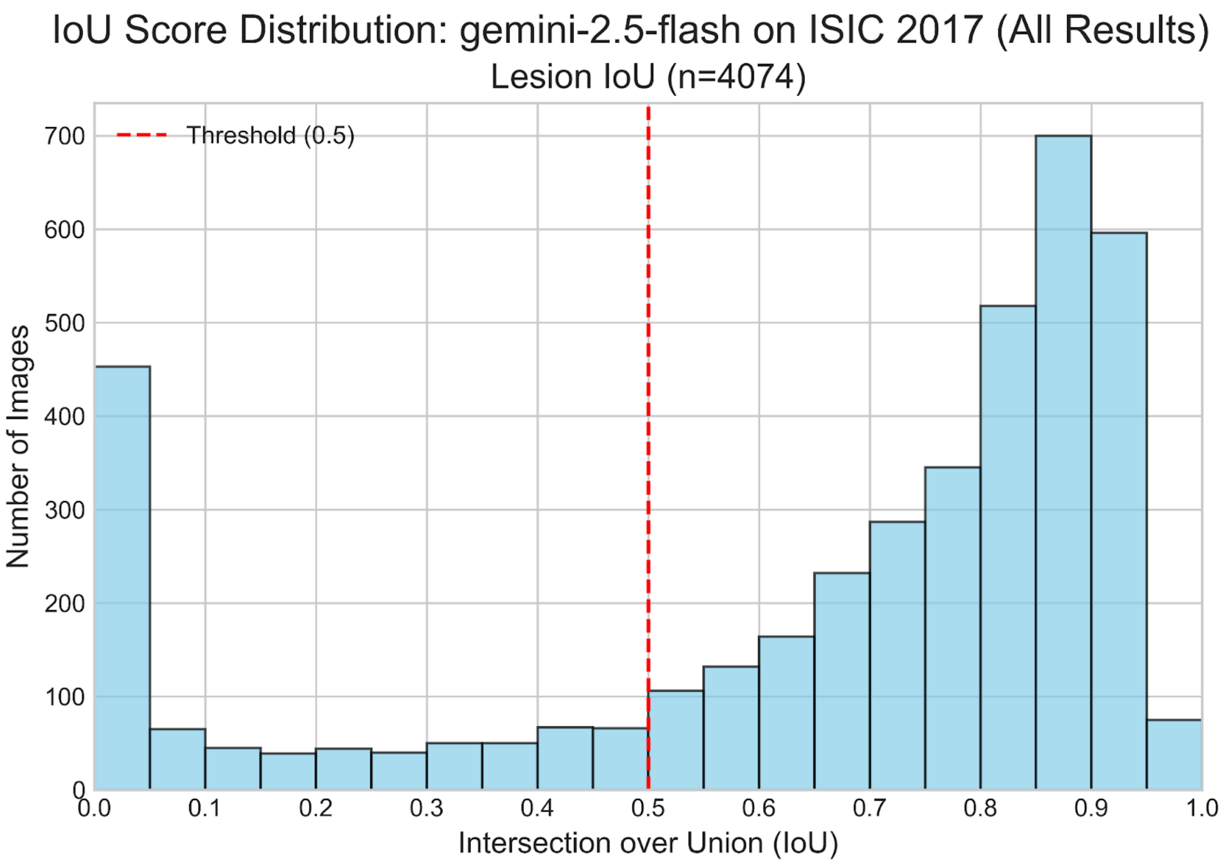
